# Loss to Follow-Up Among Patients with Kaposi Sarcoma at the Ocean Road Cancer Institute, Tanzania: A Fine–Gray Competing-Risks Analysis of Death as a Competing Event

**DOI:** 10.64898/2026.08.26.26361391

**Authors:** Emmanuel L. Lugina, Chacha Josiah Mwita, Tumaini Nyamhanga, Salum J. Lidenge, John Ngowi, Crispin Kahesa, Charles Wood, Julius Mwaiselage

## Abstract

**Purpose:** Kaposi sarcoma (KS) remains one of the most common HIV-associated malignancies in sub-Saharan Africa (SSA). While loss to follow-up (LTFU) has been well documented among KS patients managed within HIV primary care, little is known about retention after patients transition into specialized oncology care, where treatment pathways, toxicities, costs, and follow-up schedules differ substantially. Because LTFU is unlikely to occur at random, patients who disengage from care may differ systematically from those retained with respect to disease severity, treatment response, and mortality risk, potentially biasing survival estimates and underestimating cancer-related mortality. This study aimed to estimate the cumulative incidence of LTFU among patients with KS receiving care at Tanzania’s national cancer referral center, accounting for death as a competing event, and to identify factors associated with LTFU.

**Methods:** This retrospective cohort study included 251 patients with KS treated at Ocean Road Cancer Institute (ORCI) between January 2021 and December 2023. The primary outcome was LTFU, with death treated as a competing event. Cumulative incidence of LTFU at 6, 12, 18, and 24 months was estimated using the cumulative incidence function. Predictors of LTFU were assessed using univariable and multivariable Fine-Gray subdistribution hazards regression.

**Results:** Among 251 patients, 214 (85.3%) had epidemic (HIV-associated) KS and 37 (14.7%) had endemic (non-HIV-associated) KS. Males accounted for 62.2%. Accounting for death as a competing event, the cumulative incidence of LTFU was 27.6% (95% CI, 22.0–33.1) at 6 months, 36.4% (95% CI, 30.5–42.4) at 12 months, 43.3% (95% CI, 37.2–49.4) at 18 months, and 46.6% (95% CI, 40.4–52.7) at 24 months. In multivariable Fine–Gray regression, absence of oral involvement (adjusted subdistribution hazard ratio [aSHR], 0.37), reachable telephone contact (aSHR, 0.54), and initial chemotherapy rather than radiotherapy (aSHR, 0.44) were independently associated with lower risk of LTFU.

**Conclusion:** Nearly half of patients with KS were LTFU within two years, substantially limiting reliable assessment of cancer outcomes in this setting. Strengthening retention strategies, including maintaining reliable patient contact information and implementing routine phone-based follow-up, may offer feasible, scalable approaches to improve continuity of care, enhance survival monitoring, and strengthen cancer surveillance in resource-limited settings. **Keywords:** Kaposi sarcoma; loss to follow-up; cancer survival; competing risks; Fine–Gray regression; Tanzania.

## INTRODUCTION

Kaposi sarcoma (KS) remains one of the most common malignancies in sub-Saharan Africa, driven by high endemic seroprevalence of human herpesvirus-8 (HHV-8) and the ongoing HIV epidemic(1). In Tanzania, KS remains among the leading cancers affecting men and represents a substantial component of the caseload at the Ocean Road Cancer Institute (ORCI), the country’s national cancer referral center(2)Despite substantial scaling up of antiretroviral therapy (ART) coverage over the past two decades, KS continues to present at advanced stages in many sub-Saharan African (SSA) countries, with outcomes adversely affected by disease severity at diagnosis and interruptions in sustained engagement with HIV and cancer care(3).

Loss to follow-up (LTFU) has been repeatedly identified as a major challenge to the accurate assessment and improvement of KS outcomes in SSA. In a five-country analysis from the International Epidemiologic Databases to Evaluate AIDS (IeDEA) collaboration, involving 1,328 patients with newly diagnosed KS receiving care through HIV clinics, the cumulative incidence of LTFU reached 36% at 12 months and 45% at 24 months, with death treated as a competing event(3). Comparative data from South Africa, Malawi, and Zambia demonstrate substantial variation in LTFU among patients with AIDS-related KS. In a multicountry comparison, one-year LTFU was 15% in South Africa, 24% in Malawi, and 40% in Zambia. Notably, 90% of patients in South Africa were referred to an oncology department for KS care. In contrast, most patients in Malawi and Zambia were managed within HIV clinics, suggesting that differences in models of care may contribute to variation in retention and outcomes. However, the study was small and observational, and the independent effect of oncology referral on LTFU could not be established(4). Programs integrating protocol-guided chemotherapy with ART delivery and structured psychosocial support have reported high retention and favorable survival among patients with HIV-associated KS. In rural Malawi, an integrated KS care model combined standardized chemotherapy and ART with transportation reimbursement for patients with mobility-limiting disease, routine home-based psychosocial support by community health workers, adherence counseling, and active linkage to health facilities when interim illnesses were identified. At 12 months, 77% of patients were alive and retained in care, while only 5% were lost to follow-up. These findings highlight the potential importance of care-delivery models and modifiable health-system and patient-support interventions in sustaining engagement with KS care, alongside disease-related factors(5).

Most published evidence on retention in care among patients with KS in SSA derives from HIV/ART programs (3, 5–8), while substantially less is known about retention after patients enter specialized oncology services. Oncology-based KS care may involve distinct patterns of healthcare utilization, including longer travel to national or regional referral centers, repeated hospital visits, direct and indirect treatment-related costs, and the administration of systemic chemotherapy requiring toxicity monitoring in addition to ongoing ART. Commonly used KS regimens, including vincristine-based chemotherapy and taxane-based therapy such as paclitaxel, introduce treatment-related toxicities and monitoring requirements that differ from those encountered with ART alone. These differences in care pathways and treatment burden may influence both the magnitude and determinants of LTFU. However, data specifically describing retention and predictors of LTFU among patients receiving KS care in routine oncology settings in SSA remain limited.

KS remains associated with substantial mortality in SSA, although survival varies considerably according to KS subtype, disease stage, HIV status, treatment access, and healthcare setting(1). Studies from SSA have reported considerably poorer outcomes than those observed in contemporary European and North American cohorts, where survival has improved substantially over time. In Mozambique, for example, outcomes among patients with HIV-associated KS treated at a specialized center remained suboptimal despite access to cancer-directed therapy and antiretroviral treatment (9). These disparities are likely influenced by differences in stage at diagnosis, access to effective cancer and HIV treatment, supportive care, and continuity of care. Furthermore, high rates of LTFU in SSA KS cohorts may both compromise clinical outcomes through interruption of care and lead to under-ascertainment of subsequent deaths, thereby biasing estimates of survival and mortality (3).

Because LTFU is unlikely to be independent of patients’ clinical and sociodemographic characteristics, patients who disengage from care may differ systematically from those retained in care with respect to disease severity, treatment response, and underlying mortality risk. Consequently, failure to adequately account for LTFU may introduce informative censoring and lead to biased estimates of survival and cancer-related mortality (19).

This study aimed to estimate the cumulative incidence of LTFU, accounting for death as a competing event, among patients with KS receiving care at Tanzania’s national cancer referral center, and to identify factors associated with LTFU.

## MATERIALS AND METHODS

### Study design and subjects

This was a retrospective, hospital-based study in which records of all histologically confirmed KS patients who attended the Ocean Road Cancer Institute (ORCI) between January 2021 and December 2023 were reviewed. ORCI is Tanzania’s national public cancer referral center, located in Dar es Salaam, Tanzania, receiving patients with cancer from across the country and neighboring countries in Eastern, Southern, and Central Africa. A standardized data extraction form was used to retrieve patients’ information, including age, sex, marital status, religion, occupation, and place of residence at KS diagnosis, and HIV serostatus. Clinical information of the patients was also abstracted from the electronic patient medical records.

Time zero for the analysis was the date of registration at ORCI. The primary outcome was LTFU. The LTFU was defined as failure to return for care within 6 months of the last scheduled clinic visit with no evidence of death or transfer out. Time to LTFU was measured in months from ORCI registration to LTFU, and data were collected retrospectively from patients’ medical records. Patients were followed until death, and for those not known to be dead, the last known date known to be alive was either from a clinic visit, transfer to another facility, or LTFU. Active telephone tracing was undertaken where feasible to improve ascertainment of vital status among patients initially classified as LTFU, thereby minimizing misclassification bias. Patients who had discontinued care and could not be reached by telephone, despite attempts to contact them or their recorded next of kin, were classified as lost to follow-up (LTFU).

Patients who died before experiencing LTFU were classified as having experienced a competing event. Patients who were alive and retained in care at the time of last contact or who were formally transferred to another health facility for continued cancer care were treated as right-censored at the date of last confirmed follow-up.

A secondary outcome was the timing of LTFU, categorized as early or late. Early LTFU was defined as discontinuation of care within 6 months of a scheduled clinic visit. In contrast, late LTFU was defined as discontinuation occurring more than 6 months after a scheduled clinic visit. Patients who remained in care throughout follow-up were not included in these categories. Ethical approval was obtained from the Institutional Review Board of the Muhimbili University of Health and Allied Sciences (MUHAS).

### Statistical analysis

All data abstraction forms were reviewed for completeness before entry into a Microsoft Excel database. Statistical analyses were performed using R (version 4.4.2) within RStudio (version 2026.1.1.403). Continuous variables were assessed for normality using the Shapiro–Wilk test. Normally distributed variables were summarized as means with standard deviations (SD), while non-normally distributed variables were summarized as medians with interquartile ranges (IQR). Categorical variables were described using frequencies and percentages. Comparisons between groups were performed using the Student’s t-test for normally distributed continuous variables and the Mann–Whitney U test for non-normally distributed variables. Comparisons between categorical variables were performed using the chi-square test.

LTFU rates were initially estimated using the Kaplan–Meier method, and univariable comparisons were explored using the log-rank test.

Given that death precluded subsequent LTFU and was common in this cohort, the Cumulative Incidence Function (CIF) was used in the primary analysis. The cumulative incidence function (CIF) for loss to follow-up (LTFU), accounting for death as a competing event, was estimated at 6, 12, 18, and 24 months using the ‘cuminc()’ function from the cmprsk package in R. The cumulative incidence of death was also estimated over the same follow-up intervals. Median time to LTFU was reported when estimable.

Predictors of LTFU in the presence of death as a competing event were evaluated using univariate Fine and Gray sub-distribution hazards regression models. Results were expressed as sub-distribution hazard ratios (sHRs) with corresponding 95% confidence intervals (CIs). Covariates were selected based on clinical relevance and data completeness, including age, sex, disease stage, mouth involvement, initial treatment modality, treatment response, CD4 cell count, education level, marital status, residence, occupation, comorbidities, and religion. Variables with moderate missingness (<20%) were handled using multiple imputations using the ‘mice’ package. A total of five imputed datasets were generated using Predictive Mean Matching for continuous variables. Each dataset was analyzed separately, and results were pooled using Rubin’s rules. Variables with >20% missing values were excluded from the analysis.

Variables with biological plausibility and those with the outcome at p-value ≤ 0.2 in the univariate Fine Gray Model were included in the multivariate Fine Gray Model. Sub-distribution hazard ratios (SHRs) with 95% confidence intervals were reported for all covariates. Proportional subdistribution hazard assumptions were evaluated graphically.

A two-sided p-value <0.05 was considered statistically significant.

## RESULTS

We analyzed data from 251 adults with KS in this study, with a median follow-up of 13 months. Patients LTFU were slightly younger (median age 42 vs 45 years; p = 0.038). No statistically significant differences were observed for sex, religion, occupation, marital status, insurance status, education, smoking, alcohol use, or residence between patients LTFU and retained in care (Table 1).

**Table 1:** Social demographic characteristics of participants.

| Variable |  | *Entire cohort*<br>*N=251* | *LTFU* |  | *P-value* |
| --- | --- | --- | --- | --- | --- |
|  |  |  | *No*<br>*n=90* | *Yes*<br>*n=161* |  |
| *Age (Years)* | *Median (IQR)* | 43(37-52) | 45(37-57) | 42(36-49) | 0.054 |
| *Insurance* | *Yes* | 50 (19.9) | 24 (26.7) | 26 (16.1) | 0.07 |
|  | *No* | 201 (80.1) | 66 (73.3) | 135 (83.9) |  |
| *Religion* | *Christian* | 139(55.4) | 45 (50.0) | 94 (58.4) | 0.23 |
|  | *Moslem* | 112 (44.6) | 45 (50.0) | 67 (41.6) |  |
| *Referral* | *Self* | 132 (52.6) | 47 (52.8) | 85 (52.8) | 0.93 |
|  | *By hospital* | 119 (46.4) | 43 (47.2) | 76 (47.2) |  |
| *Gender* | *Male* | 156(62.2) | 60(66.7) | 96(59.6) | 0.27 |
|  | *Female* | 95(37.8) | 30(33.3) | 65(40.4) |  |
| *Occupation* | *Peasants* | 109(43.4) | 40(44.4) | 69(42.9) | 0.80 |
|  | *Non-peasants* | 142(56.6) | 50(55.6) | 92(57.1) |  |
| *Marital status* | *Married* | 139(55.6) | 51(56.7) | 88(54.7) | 0.76 |
|  | *Not married* | 112(44.4) | 39(43.3) | 73(45.3) |  |
| *Education* | *Primary and less* | 175(69.7) | 64(71.6) | 111(68.9) | 0.72 |
|  | *Secondary and above* | 76(30.3) | 26(28.9) | 50(31.1) |  |
| **Smoking** | **Yes** | 41(16.4) | 15(16.9) | 26(16.1) | 0.93 |
|  | **No** | 210(83.6) | 75(83.9) | 135(83.8) |  |
| **Alcohol** | **Yes** | 97(38.8) | 30(33.3) | 67(41.9) | 0.32 |
|  | **No** | 154(61.2) | 60(65.7) | 94(58.1) |  |
| **Residence** | **DSM** | 111(44.2) | 40(44.1) | 71(44.1) | 0.958 |
|  | **Non -DSM** | 140(55.8) | 50(55.9) | 90(55.9) |  |

*IQR- Interquartile range*

Among 251 patients with KS, 241 (85.3%) had EpKS and 37 (14.7%) had EnKS. Clinical characteristics and treatment factors were broadly similar between patients LTFU and those retained in cancer care. HIV positivity was higher among LTFU patients (77.8% vs 89.4%; p = 0.012). Symptom duration, Karnofsky Performance Status (KPS), CD4 count, hemoglobin level, disease stage, treatment modality, and response to therapy did not differ significantly between the two groups (Table 2).

**Table 2:** Clinical characteristics of participants.

| Variable |  | Entire cohort<br>N=251 | LTFU |  | P-value |
| --- | --- | --- | --- | --- | --- |
|  |  |  | No<br>N=90 | Yes<br>n=161 |  |
| Symptoms<br>Duration | Median (IQR)<br>(Months) | 12(6-12) | 12(6-12) | 12(7-24) | 0.067 |
| KPS | Median (IQR) | 90(80-90) | 90(80-90) | 90(80-90) | 0.51 |
| Hypertension | Yes | 13(5.2) | 9(10.0) | 9(5.5) | 0.34 |
|  | No | 230(91.6) | 81(90.0) | 152(94.5) |  |
| DM | Yes | 4(1.6) | 3(2.9) | 4(2.2) | 0.95 |
|  | No | 239(95.2) | 87(97.9) | 157(97.8) |  |
| HIV | Positive | 241(85.3) | 70(77.8) | 144(89.4) | 0.012 |
|  | Negative | 37(14.7) | 20(22.2) | 17(10.6) |  |
| Hb level (g/dl) | Mean ±SD | 10.2±2.5 | 10.6±2.6 | 10.1±2.4 | 0.124 |
| CD4 count | Median (IQR) | 250(128-352) | 250(104-328) | 246(146-369) | 0.472 |
| Lesion<br>number | Single | 46(18.2) | 13(14.9) | 32(20.0) | 0.43 |
|  | Multiple | 205(81.8) | 77(85.1) | 129(80.0) |  |
| Limbs<br>affected | Yes | 196(78.1) | 74(82.0) | 122(75.8) | 0.36 |
|  | No | 55(29.1) | 16(18.0) | 39(24.2) |  |
| Mouth<br>affected | Yes | 37(14.7) | 13(15.0) | 24(14.6) | 0.98 |
|  | No | 214(85.3) | 77(84.9) | 136(85.4) |  |
| Viscera<br>affected | Yes | 38(15.3) | 13(14.9) | 25(15.5) | 0.99 |
|  | No | 213(84.7) | 77(85.1) | 136(84.4) |  |
| Disseminated<br>disease | Yes | 79(31.3) | 31(34.0) | 48(29.8) | 0.61 |
|  | No | 172(68.7) | 59(66.0) | 113(70.1) |  |
| Stage | Non-locally<br>advanced | 118(47.0) | 41(45.6) | 77(47.8) | 0.757 |
|  | Locally<br>advanced | 68(27.0) | 26(29.3) | 41(25.7) |  |
|  | Disseminated | 65(25.9) | 23(26.1) | 42(26.5) |  |
| Stage | Low risk | 93(43.5) | 28(40.60) | 65(46.4) | 0.80 |
| **(EpKS only)** | **High risk** | 116(54.2) | 41(59.4) | 75(53.6) |  |
|  | **Missing** | 5(2.3) |  |  |  |
| **Treatment waiting time (Days)** | **Median (IQR)** | 9(5-29) | 8(5-27) | 9.5(5-24) | 0.823 |
| **Initial treatment** | **EBRT** | 159(59.8) | 53(63.9) | 97(66.9) | 0.641 |
|  | **Chemotherapy** | 78(31.1) | 30(36.1) | 48(33.1) |  |
|  | **No Rx** | 23(9.2) |  |  |  |
| **Treatment Response (%)** | **Median (IQR)** | 80(50-90) | 80(50-100) | 70(40-90) | 0.128 |

*IQR- Interquartile range; DM- Diabetes Mellitus; Hb – Hemoglobin; KPS-Karnofsky Performance Status; EBRT-External beam radiotherapy*

The Kaplan–Meier analysis across the imputed datasets showed a progressive increase in pooled cumulative incidence of LTFU over time. Specifically, the pooled estimated cumulative incidence of LTFU was 28.9% (95% CI: 20.9–30.7) at 6 months, 39.2% (95% CI: 28.8–49.2) at 12 months, 47.3% (95% CI: 34.9–59.0) at 18 months, and 51.4% (95% CI: 37.7–65.1) at 24 months. The median time to LTFU was 21.7 months (Fig2).

**Fig 1:**
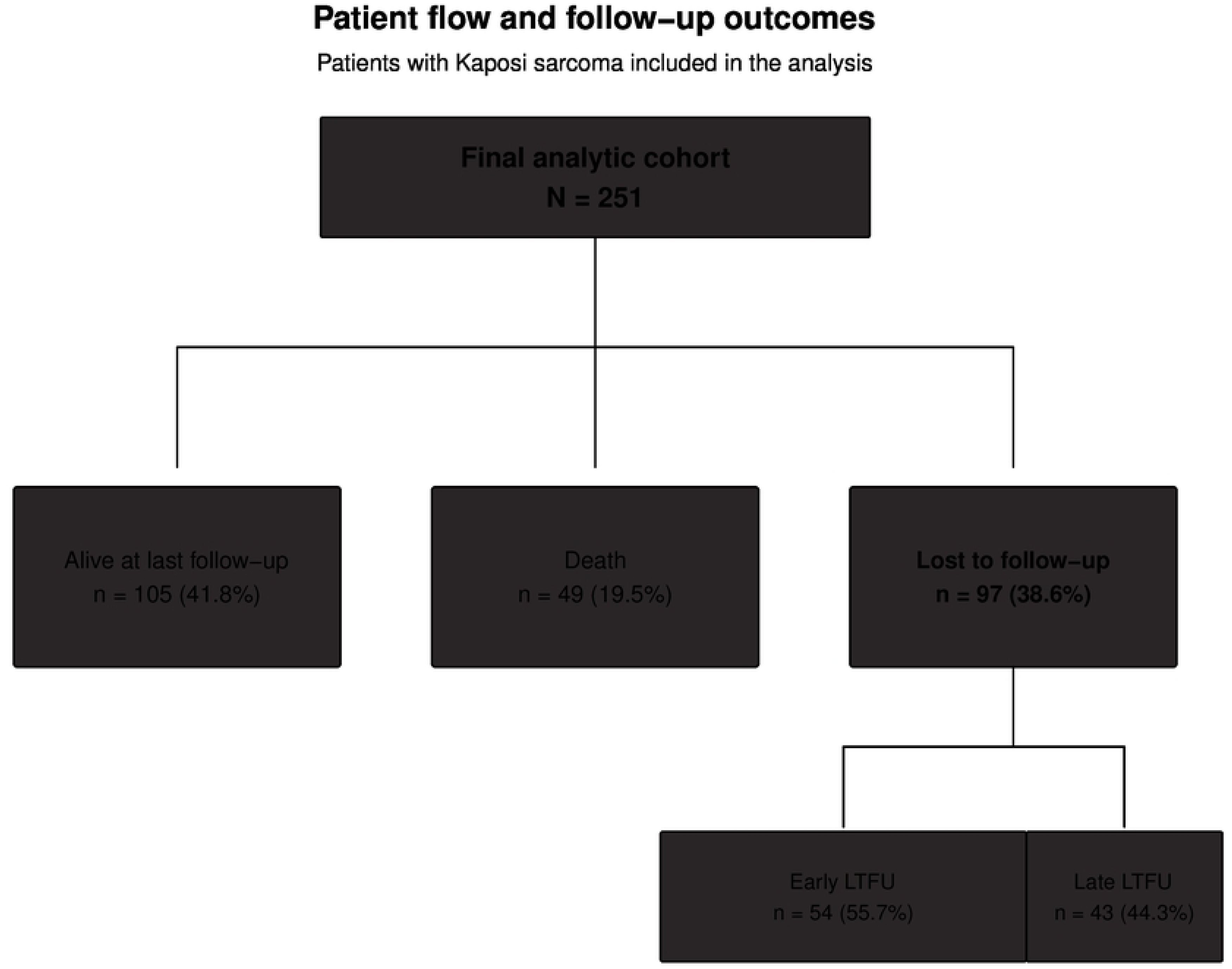
Patient flow and follow-up status among patients with KS included in the study. Among 251 patients, 105 (41.8%) were alive at last documented follow-up, 49 (19.5%) had died, and 97 (38.6%) were LTFU. Among patients who were LTFU, 54 (55.7%) experienced early LTFU and 43 (44.3%) experienced late LTFU (Fig1).

**Fig 2:**
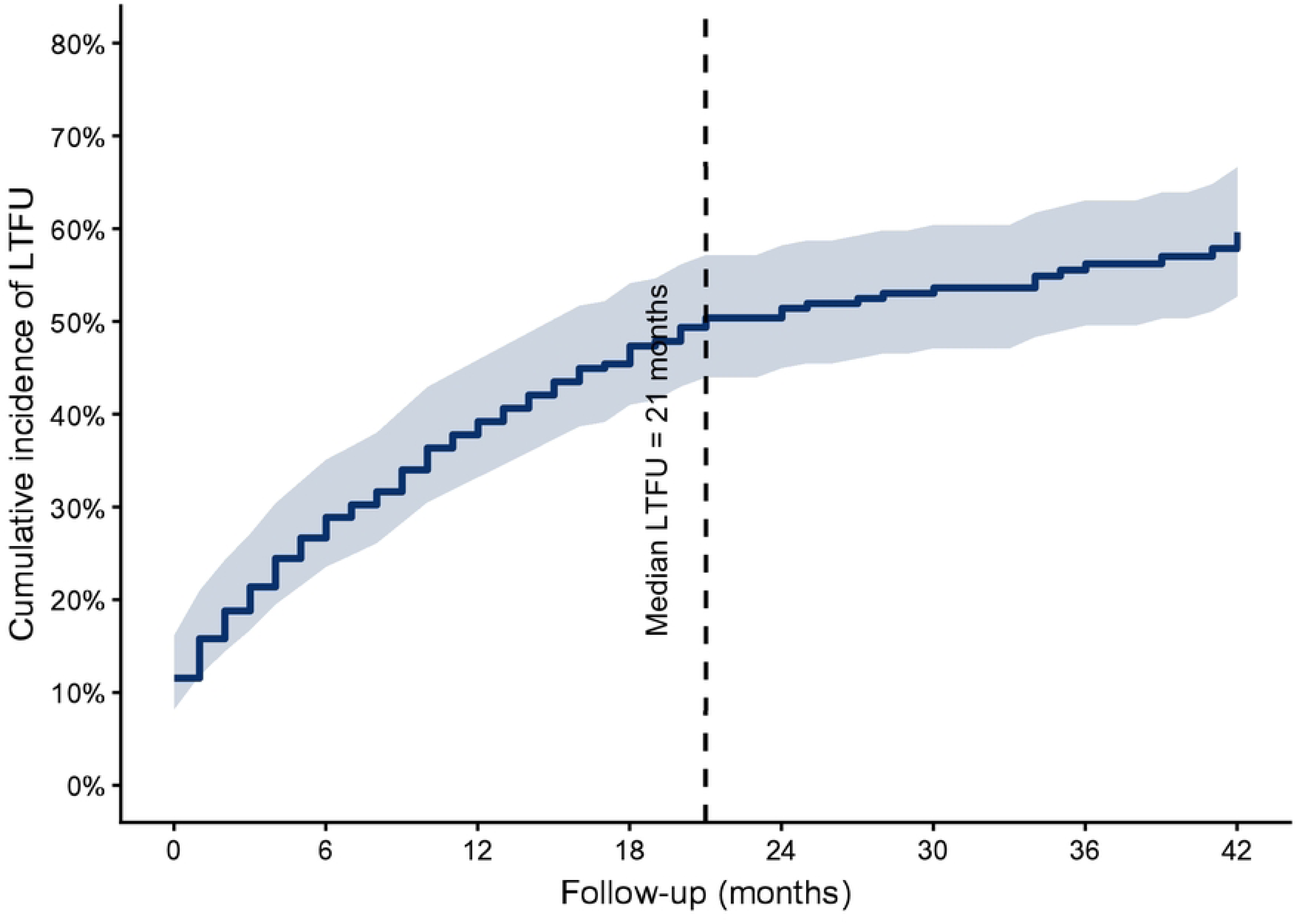
Pooled Cumulative incidence function (CIF) of LTFU based on Kaplan-Meier analysis.

Accounting for death as a competing risk, the pooled cumulative incidence of LTFU (red curve) was 27.6% (95% CI: 22.0–33.1) at 6 months, 36.4% (30.5–42.4) at 12 months, 43.3% (37.2–49.4) at 18 months, and 46.6% (40.4–52.7) at 24 months. Over the same period, the cumulative incidence of death (blue curve) was 8.8% (5.3–12.3), 11.8% (7.8–15.8), 13.2% (9.0–17.4), and 14.4% (10.1–18.8), respectively (Fig3).

**Fig 3:**
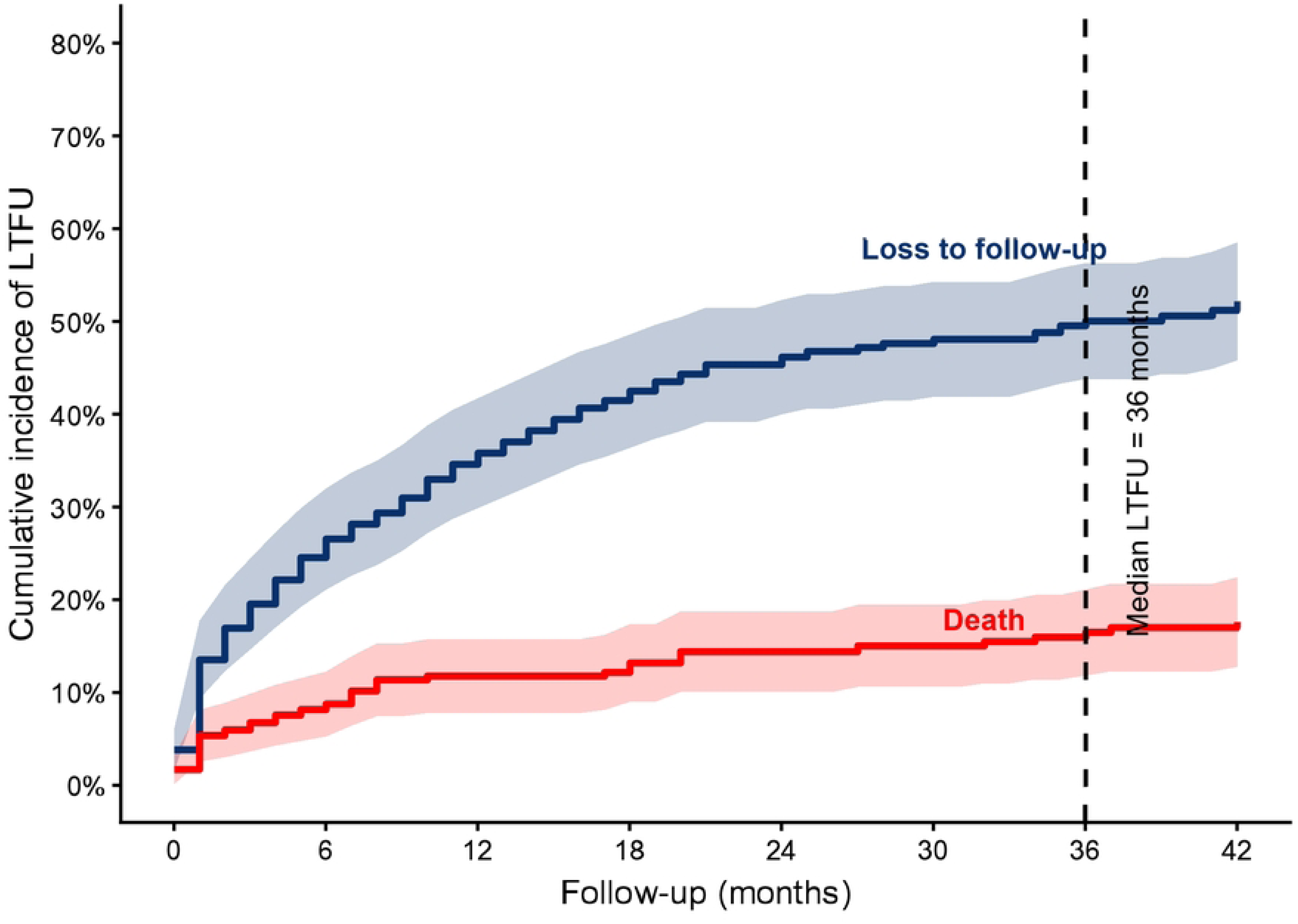
Pooled Cumulative incidence function (CIF) of LTFU with death as a competing event among KS patients.

In univariable Fine–Gray competing-risk regression, phone non-reachability was significantly associated with an increased subdistribution hazard of loss to follow-up (LTFU). Patients whose phone contacts were unreachable had a higher cumulative incidence of LTFU than those whose contacts were reachable (sHR, 1.74; 95% CI, 1.02–2.98; p=0.025). Other evaluated covariates were not significantly associated with LTFU in univariable Fine–Gray competing-risk models.

In the multivariable Fine–Gray competing-risk regression model, oral Kaposi sarcoma involvement and phone non-reachability were independently associated with LTFU. Patients with oral KS involvement had a significantly higher cumulative incidence of LTFU compared with those without oral involvement (adjusted subdistribution hazard ratio [aSHR], 1.85; 95% CI, 1.12–3.07; p=0.017). Similarly, patients whose phone contacts were not reachable had an increased subdistribution hazard of LTFU (aSHR, 1.84; 95% CI, 1.05–3.21; p=0.017).

Initial treatment modality was also associated with LTFU. Patients who received chemotherapy had a lower cumulative incidence of LTFU compared with those initially treated with radiotherapy (aSHR, 0.67; 95% CI, 0.45–0.99; p=0.045). This finding should be interpreted cautiously, as treatment allocation may have been influenced by underlying disease characteristics and clinical factors.

Age, HIV status, and treatment response category were not significantly associated with LTFU in the adjusted model. Responders had a similar cumulative incidence of LTFU compared with non-responders (aSHR, 0.87; 95% CI, 0.48–1.61; p=0.628) (Table 3).

**Table 3:** Univariable and multivariable Fine–Gray competing-risks regression analysis of factors associated with loss to follow-up among patients with Kaposi sarcoma, with death treated as a competing event.

| *Variable* |  | *Univariable analysis* |  |  | *Multivariable analysis* |  |  |
| --- | --- | --- | --- | --- | --- | --- | --- |
|  |  | *p-value* | *sHR* | *95% CI* | *p-value* | *sHR* | *95% CI* |
| *Age* |  | 0.15 | 0.99 | 0.99–1.003 | 0.24 | 0.99 | 0.98–1.01 |
| *Sex* | *Male* | 1 |  |  |  |  |  |
|  | *Female* | 0.28 | 1.19 | 0.87–1.62 |  |  |  |
| *Alcohol use* | *No* | 1 |  |  |  |  |  |
|  | *Yes* | 0.15 | 1.31 | 0.91–1.88 |  |  |  |
| *Insurance* | *No* | 1 |  |  |  |  |  |
|  | *Yes* | 0.69 | 0.92 | 0.61–1.39 |  |  |  |
| *HIV* | *Negative* | 1 |  |  | 1 |  |  |
|  | *Positive* | 0.27 | 1.33 | 0.79–2.22 | 0.55 | 1.19 | 0.65–2.20 |
| *Oral KS* | *No* | 1 |  |  | 1 |  |  |
|  | *Yes* | 0.19 | 1.37 | 0.85–2.18 | 0.017 | 1.85 | 1.12–3.07 |
| *Hemoglobin level (g/dl)* |  | 0.25 | 0.96 | 0.90–1.03 |  |  |  |
| *Symptom duration (Months)* |  | 0.09 | 0.99 | 0.98–1.002 |  |  |  |
| *CD4 count* |  | 0.102 | 1.00 | 0.99–1.003 |  |  |  |
| *Duration of HAART use* |  | 0.68 | 0.99 | 0.99–1.005 |  |  |  |
| *Stage* | *1* | 1 |  |  |  |  |  |
|  | *2* | 0.34 | 0.85 | 0.58–1.23 |  |  |  |
|  | *3* | 0.86 | 1.04 | 0.69–1.55 |  |  |  |
| *Initial treatment* | *RT* | 1 |  |  |  |  |  |
|  | *CT* | 0.42 | 0.86 | 0.59–1.24 | 0.04 | 0.67 | 0.46–0.99 |
| *Phone not reachable* | *No* | 1 |  |  | 1 |  |  |
|  | *Yes* | 0.02 | 1.74 | 1.02–2.98 | 0.017 | 1.84 | 1.95–3.21 |
| *Treatment response* | *No* | 1 |  |  |  |  |  |
|  | *Yes* | 0.68 | 0.89 | 0.47–1.68 | 0.63 | 0.87 | 0.48–1.61 |

*sHR*- subdistribution hazard ratio; *aSHR*- adjusted subdistribution hazard ratio; *EBRT*- external beam radiotherapy; *CT*- chemotherapy; *KS*- Kaposi sarcoma; *HAART*- highly active antiretroviral therapy; *PNR*-Phone not reachable

The model was derived using multivariable Fine–Gray competing-risk regression, with death treated as a competing event. Squares represent adjusted subdistribution hazard ratios (aSHRs), and horizontal lines represent 95% confidence intervals. The vertical reference line at an aSHR of 1 indicates no association with loss to follow-up. Values greater than 1 indicate an increased cumulative incidence of loss to follow-up, whereas values less than 1 indicate a reduced cumulative incidence. Oral KS involvement and phone non-reachability were independently associated with a higher cumulative incidence of loss to follow-up. In contrast, initial chemotherapy treatment was associated with a lower cumulative incidence of loss to follow-up than radiotherapy. Age, HIV status, and treatment response were not significantly associated with loss to follow-up 9 (Fig4).

**Fig 4.**
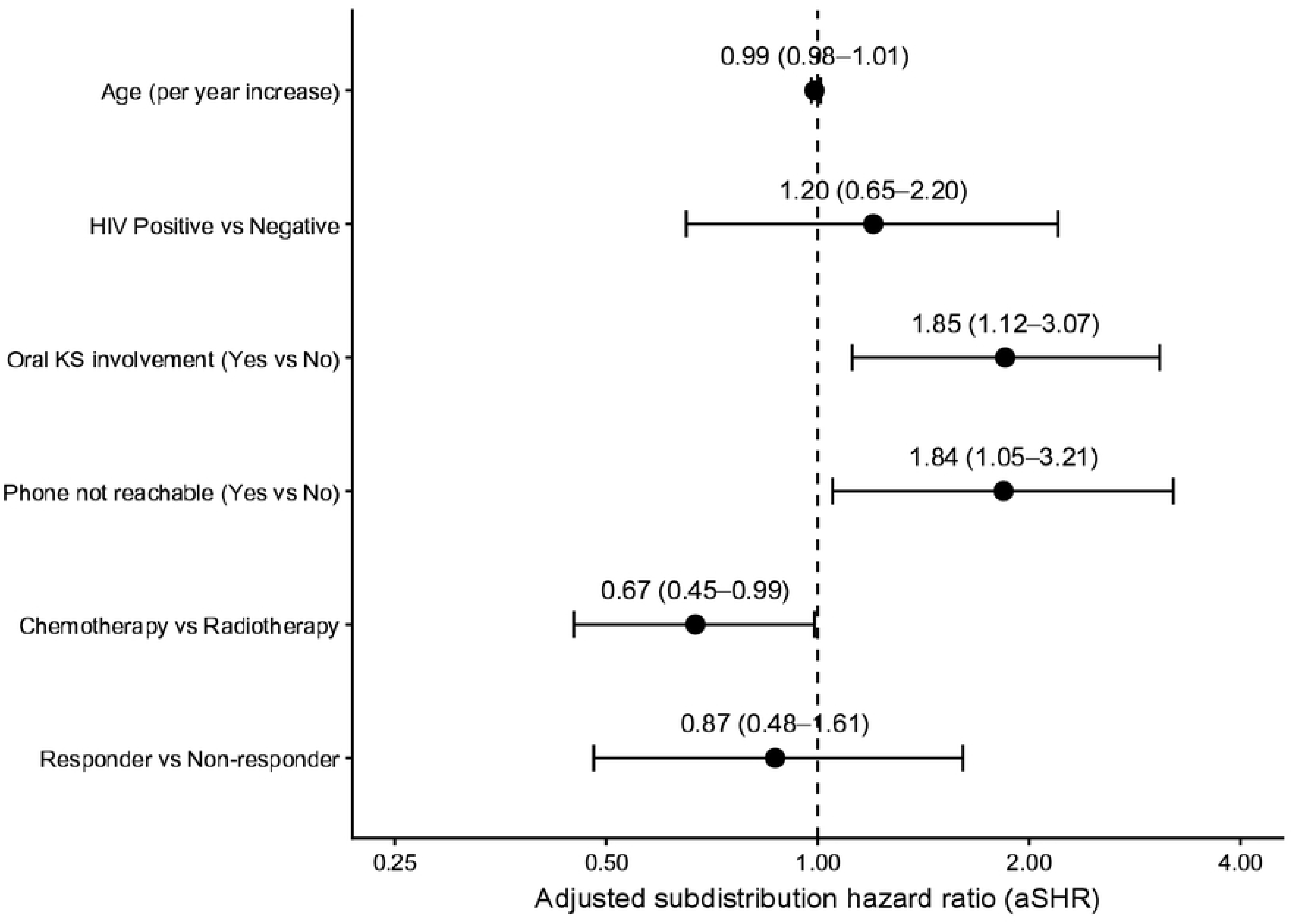
Forest plot of adjusted subdistribution hazard ratios for predictors of loss to follow-up among patients with Kaposi sarcoma. Death was treated as a competing event.

Among patients who experienced loss to follow-up, 141 (56.2%) were classified as having early LTFU and 110 (43.8%) as having late LTFU. Age distribution was similar between patients with early and late LTFU (median age 44 vs 42 years, respectively; p=0.16). However, baseline performance status differed significantly between groups, with patients experiencing late LTFU having a higher mean KPS compared with those with early LTFU (87±8 vs 85±10; p=0.04) (Table 4).

**Table 4:**
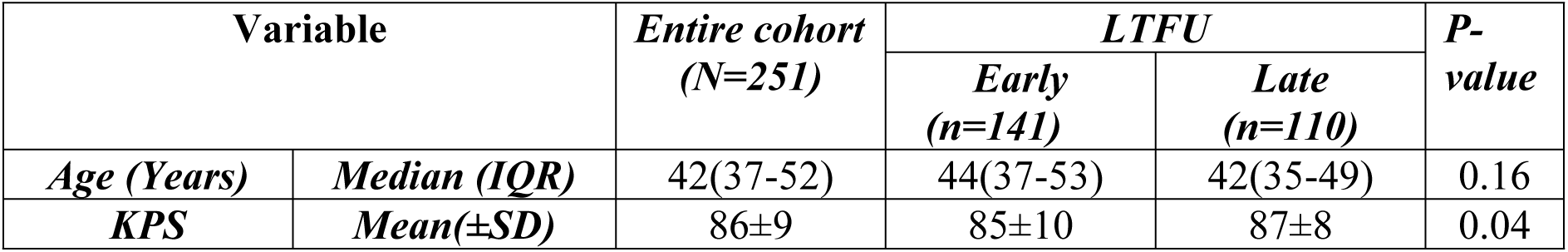
Characteristics associated with timing of loss to follow-up among patients with KS who experienced LTFU.

| Variable |  | *Entire cohort*<br>*(N=251)* | *LTFU* |  | *P-value* |
| --- | --- | --- | --- | --- | --- |
|  |  |  | *Early*<br>*(n=141)* | *Late*<br>*(n=110)* |  |
| *Age (Years)* | *Median (IQR)* | 42(37-52) | 44(37-53) | 42(35-49) | 0.16 |
| *KPS* | *Mean(±SD)* | $86\pm 9$ | $85\pm 10$ | $87\pm 8$ | 0.04 |

## Discussion

To our knowledge, this is among the first studies in SSA to formally quantify LTFU as a primary outcome among patients with KS receiving care in a dedicated oncology referral setting, using a competing-risks framework that accounts for death as a competing event.

In this analysis, the cumulative incidence of LTFU remained high over time, reaching 34.6% and 46.6% at 12 and 24 months, respectively, when accounting for death as a competing risk. These findings underscore the substantial disengagement from care in oncology settings among patients with KS, particularly in LMICs. The use of a competing-risk framework is methodologically important, as failure to account for death may overestimate LTFU or misclassify mortality as disengagement from care, as demonstrated by the higher estimates obtained with the Kaplan–Meier method in this study.

The substantial LTFU observed in this study is consistent with findings from other studies in SSA. This magnitude is remarkably similar to that reported in a multicountry SSA cohort of patients with KS, in which the cumulative incidence of LTFU was 36% at 1 year and 45% at 2 years. Importantly, that study also accounted for death as a competing event, making it the most methodologically comparable study to the present analysis. Despite the similarity in estimates, the care settings differed substantially: the multicountry cohort comprised patients identified within HIV primary-care programs, whereas our patients were managed within a national cancer referral center. The comparable magnitude of LTFU therefore suggests that loss from longitudinal KS care may remain substantial even when care is delivered within a specialist oncology setting (10)

Our findings are also broadly consistent with evidence from a South African tertiary medical oncology service, in which patient retention was limited. One-year survival and retention were 60.4% and 72.3% in the earlier and later ART eras, respectively, while two-year survival and retention were 39.8% and 66.7%. However, these estimates represent the combined outcome of survival and retention and therefore cannot be interpreted as direct estimates of LTFU. Nevertheless, the study demonstrates that attrition from oncology care remains a significant problem even within specialist cancer services(6). Evidence from Tanzania also suggests substantial difficulties with follow-up among patients with KS. Chalya et al., in a retrospective study conducted at a tertiary hospital, reported that among patients who survived the initial treatment period, only 33% were available for 12-month follow-up, with 67% classified as lost to follow-up. However, this study was primarily designed to evaluate treatment outcomes and survival rather than LTFU, and follow-up was limited to 12 months. Furthermore, LTFU was assessed as an observed end-of-follow-up status rather than as a time-to-event outcome accounting for competing mortality. Its estimate therefore should not be directly compared with the cumulative incidence in our study. Nevertheless, the finding provides important local evidence that poor continuity of care has been a longstanding problem among Tanzanian patients with KS(11). Taken together, the available evidence indicates that LTFU is a substantial problem among patients with KS in SSA. However, estimates vary depending on the definition of LTFU, the duration of follow-up, the clinical setting, and the analytical approach. The similarity between our 1- and 2-year cumulative incidence estimates and those from the multicountry IeDEA cohort is particularly notable despite differences in healthcare setting. In contrast, the higher observed attrition reported by Chalya et al. may partly reflect differences in the definition and ascertainment of LTFU, as well as the absence of a competing-risks framework. Our findings therefore provide setting-specific evidence that substantial LTFU persists among KS patients receiving care within a specialist oncology service.

The high incidence of LTFU observed in this study may reflect a combination of patient-, treatment- and health-system-level barriers associated with prolonged cancer care in a national referral setting. Patients referred to a national cancer center may travel considerable distances and incur substantial direct and indirect costs associated with repeated hospital attendance. In addition, KS treatment may require multiple clinical encounters for chemotherapy, radiotherapy, supportive care and subsequent surveillance. After completion of active treatment, the perceived benefit of continued hospital attendance may decline, potentially increasing the risk of disengagement from follow-up. However, these explanations remain hypotheses because socioeconomic barriers, distance from the cancer center, reasons for discontinuing care, and transfers to other facilities were not systematically captured in our dataset. Importantly, the association between an unreachable telephone number and LTFU provides empirical support for the role of communication continuity in retention. This finding suggests that interventions such as verifying contact information, sending appointment reminders, and systematically tracing patients who miss scheduled visits may be feasible strategies to reduce LTFU. Further research is needed to explore the specific barriers and facilitators influencing retention in oncology care among patients with Kaposi sarcoma.

In the multivariate Fine-Gray model in this study, patients without oral involvement had a 63% lower risk of LTFU than those with oral involvement. Oral involvement in Kaposi’s sarcoma has been linked to more advanced disease, malnutrition as a result of poor oral intake, and poorer functional status, which may adversely affect retention in care(12, 13). Patients with reachable telephone contacts (higher notification/traceability rates) had a 46% lower risk of LTFU, underscoring the critical role of basic communication infrastructure in maintaining continuity of care and facilitating patient tracing(14). In this context, accessible phone contact serves as a practical link between patients and the healthcare system, enabling timely follow-up, appointment reminders, and re-engagement of individuals who miss scheduled visits(14). Mobile phone tracing is more successful and cost-effective than physical tracing in reaching patients in SSA (15). This finding is particularly relevant in the context of ORCI, a national tertiary cancer center that serves patients referred from across Tanzania. Unlike patients receiving care within geographically defined primary-care settings, many ORCI patients reside outside Dar es Salaam, making routine physical tracing after missed appointments difficult and resource-intensive. The IeDEA Kaposi sarcoma tracer study demonstrated that telephone tracing can help re-establish the status of patients classified as LTFU. However, physical/community tracing was also important in their setting (7). In the ORCI context, where patients may live considerable distances from the institution, reliable telephone contact may therefore be particularly important for maintaining communication and identifying patients who miss scheduled visits. Thus, the association between unreachable telephone contact and LTFU in our study may reflect, at least in part, the challenges of maintaining patient contact across a geographically dispersed referral population.

Additionally, patients who received chemotherapy as the initial treatment modality had a 57% lower sub-distribution hazard of loss to follow-up (LTFU) than those managed with single-fraction radiotherapy, after adjustment for other covariates. This association likely reflects differences in care delivery and patient selection, as chemotherapy is typically administered through structured, longitudinal care pathways that promote sustained engagement. This schedule creates a “rhythm of care” in which patients are repeatedly seen, assessed, and counseled. Each visit is an opportunity for healthcare workers to re-verify contact info and address barriers to care. In contrast, single-fraction radiotherapy is typically delivered in a single session, which offers limited opportunity for longitudinal relationship-building. Indeed, frequent patient contact has been shown to enhance retention in oncology in SSA (16).

The cumulative incidence of death increased progressively over time, reaching 8.8% (95% CI: 5.3–12.3) at 6 months, 11.8% (7.8–15.8) at 12 months, 13.2% (9.0–17.4) at 18 months, and 14.4% (10.1–18.8) at 24 months. In contrast, the cumulative incidence of LTFU rose more steeply over the same period, indicating that patient LTFU exceeded mortality in this cohort. The observed 2-year cumulative mortality of 14.4% is lower than that reported in many SSA cohorts, where mortality frequently exceeds 20–30% at similar time points (30), but remains higher than estimates from high-income settings (17). This likely reflects a combination of improved access to ART and oncology services, alongside persistent challenges such as late-stage presentation and health system constraints. The cumulative incidence of LTFU increased more steeply than mortality, suggesting that disengagement from care represents a major competing event in this cohort. This pattern is consistent with reports from SSA, where structural barriers to care, socioeconomic constraints, and fragmentation between HIV and oncology services contribute substantially to attrition(3). Importantly, patients classified as LTFU represent a heterogeneous group, and prior studies in similar settings suggest that a substantial proportion may have died without formal documentation(18). As such, the observed mortality is likely underestimated, and the true burden of death may be considerably higher.

The occurrence of both early and late LTFU suggests that disengagement from oncology care was not confined to the initial period following diagnosis or treatment. This finding highlights the need for retention strategies that address both early engagement in care and sustained follow-up. In this study, patients experiencing early LTFU were more likely to be younger and to have poorer baseline functional status. The association with younger age may reflect underlying socioeconomic constraints, as younger patients in low-resource settings often face greater barriers to sustained engagement in care, including competing work obligations and limited financial resources for transport and clinic attendance. In contrast, poorer functional status likely reflects more advanced disease and a higher symptomatic burden at presentation. Importantly, early LTFU may, in part, represent unascertained mortality. Patients with severe disease may deteriorate rapidly and die at home before a subsequent clinic visit, leading to misclassification as early LTFU rather than death.

This study is among the first to quantify LTFU among patients with KS in an oncological care setting in SSA. A key methodological strength is the use of competing risks analysis, which appropriately accounts for death as a competing event when estimating the cumulative incidence of LTFU, thereby avoiding overestimation inherent in standard survival approaches. However, several limitations merit consideration. First, outcome misclassification is possible, as some patients classified as LTFU may in fact have died without documentation, particularly in the absence of robust active follow-up or patient tracing systems. This could result in underestimation of mortality and corresponding overestimation of LTFU. Second, LTFU may not be a non-informative process; rather, it is likely informative, with patients who have advanced disease, poor performance status, or socioeconomic barriers being more prone to disengagement from care. This introduces the potential for selection bias and may distort the observed incidence of LTFU. Consequently, the estimates should be interpreted with caution, given the possibility of both residual outcome misclassification and informative censoring. Third, the single-center design in a tertiary referral hospital setting may limit external validity, as patient characteristics, care pathways, and retention patterns may differ in peripheral or non-specialist settings.

In conclusion, this study demonstrates that substantial LTFU among patients with KS in Tanzania poses a major challenge to the validity of survival estimates and evaluation of cancer outcomes. This limitation is becoming increasingly important as access to oncologic and HIV-directed therapies expands across SSA, increasing the need for reliable longitudinal outcome monitoring. Without effective, context-specific retention strategies for KS and other HIV-associated malignancies, survival estimates will remain vulnerable to bias, and the interpretation of cancer epidemiology data in the region will remain constrained.

Our findings highlight the importance of maintaining reliable patient contact information as a practical component of retention strategies. Patients without reachable phone contacts had a higher risk of LTFU, suggesting that accessible communication channels can strengthen the link between patients and healthcare systems through appointment reminders, early identification of missed visits, and timely re-engagement of patients who disengage from care. Integrating routine phone-based follow-up into cancer care pathways may be a feasible, scalable intervention to improve retention in resource-limited settings.

## Abbreviations

KS: Kaposi sarcoma
EpKS: Epidemic Kaposi sarcoma
EnKS: Endemic Kaposi sarcoma
IRQ: Interquartile range
LTFU: Loss to follow-up
MUHAS: Muhimbili University of Health Sciences
SSA: Sub-Saharan Africa
ORCI: Ocean Road Cancer Institute

## Authors’ contributions

Conception and Design: ELL, JDM, and SJL

Administrative Support: ELL, JDM, and SJL

Collection and assembly of data: ELL

Data Analysis and Interpretation: ELL, JDM, and SJL

Manuscript Writing: ELL, JDM, and SJL

Final approval of Manuscript: All authors

Accountable for all aspects of the work: All authors

## Data Availability

The datasets used and analyzed during the current study are available from the corresponding author on reasonable request.

## Acknowledgments

We express our sincere gratitude to the administration and staff of the Ocean Road Cancer Institute for their unwavering support throughout this study. We also acknowledge the TAMTRIP-ORCI program for its substantial contribution to our research training and capacity development.

## Ethics approval and consent to participate

Ethical approval for this study was obtained from the Muhimbili University of Health and Allied Sciences Institutional Research and Ethics Committee (MUHAS IRB). The Ocean Road Cancer Institute granted permission to conduct the study. Given the retrospective nature of the study and the use of secondary data, the MUHAS IRB approved a waiver of informed consent. All data were analyzed in an anonymized and aggregated manner, and no patient-identifiable information was included.

## Consent for publication

No individually identifiable data – not applicable.

